# COVID-19 death rates by age and sex and the resulting mortality vulnerability of countries and regions in the world

**DOI:** 10.1101/2020.05.17.20097410

**Authors:** Christophe Z Guilmoto

**Affiliations:** Institut de recherche pour le Développement, CEPED Université de Paris, 45 rue des Sts-Pères F-75006 Paris

## Abstract

The growing number of series on COVID-19 deaths classified by age and sex, released by national health authorities, has allowed us to compute age and sex patterns of its mortality, based on 183,619 deaths from Western Europe and the USA. We highlight the specific age schedule of COVID-19 mortality and its pronounced excess male mortality and we then apply these COVID-19 death rates to world populations, in 2020. Our results underscore that considerable variations exist between world regions, as concerns the potential impact of COVID-19 mortality, because of their demographic structures. When compared to younger countries in Sub-Saharan Africa, the vulnerability to COVID-19 mortality is shown to be 17 times higher in several industrialized countries of East Asia and Europe. There is a high correlation (r^2^= .44) between demographic vulnerability to COVID-19 mortality and current COVID-19 death rates.

**Abstract:** COVID-19, mortality, age structures, death rates, Europe, USA.

**Background:** The data available on infection and death rates from COVID-19 have pointed to the elderly’s vulnerability to pandemics, especially elderly men. However, current models have not yet incorporated the growing volume of information on deaths by age and sex that is being released by statistical offices and health authorities. These newly available data allow us to examine the specific age and sex patterns of COVID-19 mortality and to estimate the impact of specific demographic structures on potential COVID-19 mortality worldwide.

**Methods:** We use the data available on May 15, 2020, from the nine countries with the largest series of deaths, disaggregated by age and sex: Belgium, France, Germany, Italy, Netherlands, Spain, Sweden, the UK, and the USA. Using 183,619 deaths (60.2% of all currently estimated COVID-19 deaths), we estimate the sex-specific death rates, by 5-year ranges, for two large death samples: USA and Western Europe. We compare these mortality rates with Gompertz models and with life tables of the world’s population, estimated by the United Nations. We apply these COVID-19 mortality rates by sex and 5-year group to the 2020 age and sex structures of world countries and regions, and obtain an index summarizing the relative magnitude of their potential vulnerability to COVID-19 mortality because of their demographic structures.

**Findings:** COVID-19 death rates cannot be computed below age 15. COVID-19 death rates from age 15–19 years to 90+ increase by a factor of 3 of every ten years, at a rate that is faster than general mortality. Male mortality from COVID-19 is systematically higher than female mortality, with a peak of excess male mortality occurring among 55-59-year-olds. Age and sex structures show considerable variations across countries, in terms of vulnerability to COVID-19 mortality. It transpires that the youngest countries in Central Africa are 17 times less vulnerable than aging countries, such as Japan.

**Interpretation:** Whereas the true intensity of the ongoing COVID-19 pandemic remains underestimated by existing statistics, this unique mortality dataset shows that the regularity of the distribution of COVID deaths by age and sex is in line with the standard Gompertz mortality equation; thus confirming the quality of the first death samples and the unique age and sex patterns of COVID-19 mortality.

COVID-19 death rates tend to be negligible below age 15 and cannot be used for analysis. The rate of progression of death rates by age is faster than that of general mortality. This feature places the elderly population in a particularly vulnerable situation compared to younger adults. Male excess mortality from COVID-19 also appears far more pronounced than in general mortality patterns, with men aged 40–59 years being almost 2.5 times more likely to die than women of the same age.

Our analysis also points to considerable variations between world regions, as concerns the potential impact of COVID-19 mortality, because of their demographic structures. The COVID-19 structural vulnerability index ranges from .28 in Western or Middle Africa to 2.6 in Southern Europe. There is a high correlation between demographic vulnerability to COVID-19 mortality and current COVID-19 death rates (r^2^= .44 for 188 countries).

## 1. Introduction

The first data on the infection and death rates of COVID-19 have pointed to the elderly’s vulnerability to pandemics, and in particular elderly men.^1–4^ Thus far, existing models of COVID-related mortality have relied on the death rates of infected populations and have not incorporated the growing volume of information that is being released by statistical offices and health authorities.

Since April 2020, new series about COVID-related deaths have become available; classified by date, age group and sex (see also Appendix 1 for sources).^5,6^ These series permit us to chart the age and sex patterns of pandemic mortality — even if the reported mortality levels still underestimate the true intensity of COVID-19 mortality. Using these, it is possible to estimate the impact of specific demographic structures on the potential mortality of COVID-19, worldwide and provide a detailed demographic picture of the current and potential impact of the pandemic.^7–9^

This paper begins with a description of the recent data that will be used for a disaggregated age analysis of COVID-19 mortality. We also explain how age and sex data can be standardized across countries, and how the resulting age and sex schedules of COVID-19 mortality can be applied to all countries, to assess their respective vulnerability to the pandemic. The next section presents our results: the age and sex death rates attributed to COVID-19, the sex ratio by age of COVID-19 mortality, and the potential impact of COVID-19 mortality on all countries and regions. We conclude with the lessons drawn from our analysis and the unanswered questions that this research brings to light.

## 2. Deaths

### 2.1. Countries selected for the study

We collected the latest data – available on May 15, 2020 – on COVID-19 deaths by age and sex. We limited our analysis on the 19 countries that had a death toll, attributed to COVID-19, of above 2000 deaths. Countries with smaller numbers of deaths – such as Norway, Portugal and South Korea – were not considered, as the limited size of their (number of deaths) sample did not allow testing of the consistency of death rates by age and sex.

However, we also had to exclude several countries without available disaggregated deaths by age and sex from our sample: Brazil, Iran, China, Turkey, Mexico, India, Peru and the Russian Federation. In addition, death data from Ecuador could also not be used, as there is no available disaggregation by sex. Similarly, the lack of deaths disaggregated by age and sex precluded the use of figures from Canada. A list of these countries, including the websites of the main statistical sources on COVID-19 mortality, can be found in Appendix 1.

Our final sample covered nine countries: Belgium, France, Germany, Italy, Netherlands, Spain, Sweden, England and Wales, and the USA. Together, they accounted for more than 84% of deaths presently attributed to the COVID-19 pandemic worldwide; although, death statistics disaggregated by age and sex were often not available for all of the COVID-19 deaths within these countries.

The 183,619 deaths, with age or sex information, represent 78.6% of the total deaths currently attributed to COVID-19, on the same date in the nine countries of our sample.^10^ Coverage (deaths by age and sex/all deaths) is notably poor for the USA: its sample includes only 54,860 deaths with age and sex particulars out of the 87,248 COVID-19 deaths that were estimated on the same date (i.e. 62.9 %).

This sample represents the largest available sample of COVID-19 deaths to date, and accounts for 60.2% of all deaths attributed to COVID-19 on the same date.^10^ This is in comparison to the previous samples, used for mortality estimation purposes, which relied on only a fragment of deaths, recorded in China, the USA, and Italy.^8,11,12^

### 2.2. Reclassification and computation of age and sex death rates

With the exception of England and Wales and the Netherlands, deaths were not available by sex and 5-year age group. In the remaining countries of our sample, deaths were available by sex and various age groups (mostly 10-year and 20-year age groups). For this reason, quinquennial death rates by sex had to be estimated for these countries. Appendix 2 provides details of the reclassification procedures we followed and Appendix 3 displays the COVID-19 death rates by age and sex for each country computed from the original data.

Reclassified deaths were then collected into three different datasets:

- USA
- Western Europe (Belgium, France, Germany, Italy, the Netherlands, Spain, Sweden, and the England and Wales)
- All: USA and Western Europe

Age- and sex-specific death rates were then computed after adjustment for coverage levels, using the corresponding age and sex population figures for 2020, as estimated by the United Nations. ^13^ The resulting series of death rates were then compared with the Gompertz equation, which is commonly used in adult mortality analysis.^14^

Two observations are worth noting concerning our methodology. First, age- and sex-specific death rates were tested for consistency in each country (see Appendix 3), prior to their inclusion in the database, which precluded the use of data from countries with small numbers of reported COVID deaths. Second, COVID death rates in our sample remain underestimated, as the actual death toll is still incomplete because of the ongoing pandemic combined with the under-registration of COVID-19 deaths outside of hospitals.

### 2.3. Estimation of the potential impact of age—and sex structures on COVID-19 mortality

Detailed age- and sex-specific death rates allowed us to go one step further in measuring COVID-19’s potential impact. We aimed to apply the same mortality schedule to populations with different age and sex structures, in order to simulate the impact of COVID mortality and to compare the resulting effect on overall mortality. Our procedure was as follows:

- We applied the COVID-19 death rates, derived from our sample, to the 2020 age and sex structures (United Nations data). ^13^
- We obtained a simulated number of deaths and then computed COVID-19 death rates for all national and regional populations.
- Thereafter, we computed a standardized COVID-19 structural vulnerability index (CSVI), by dividing all simulated death rates by the average world death rate.

This standardized CSVI index indicates whether specific regions or countries would be relatively more or less affected by COVID-19 mortality. By definition, CSVI took a value of one for the entire world. CSVI reached values of greater than 1 in older countries, and lower than 1 in countries with younger age structures.

While COVID death rates used herein are underestimated, this underestimation should have a limited influence on the standardized COVID-19 vulnerability index, as CSVI is a comparative ratio, based only on the age schedule of deaths by sex in our sample and not on its level. In theory, different age patterns of COVID-19 mortality, in countries not covered in our sample, may bias this index slightly. However, we ran the same simulations using the mortality patterns of the USA and Western Europe and found that the resulting differences in the level of vulnerability index were negligible (results available on request).

## 3. Findings

In this section, we first examine the distribution of COVID death rates in our sample by age and sex, and then we compare them with the general observed death rates worldwide over in 2015–20. Next, we examine the sex differentials by age and region. Finally, we evaluate the differential impact of age and sex structures on the overall level of COVID-19 mortality worldwide.

### 3.1. Age schedule of COVID-19 mortality

Figure 1 presents the COVID death rates computed for the different samples, by age for men and women (see also Appendix 4 for rates). It may be noted that, at the outset, COVID death rates tend to be negligible below age 15 in all the death samples used herein. The total number of deaths below age 15 in our sample is 14 deaths. As a result, death rates below 15 are computed on extremely rare events and are not significantly different from zero. Therefore, the rest of the analysis focuses on mortality from age 15–19 years upwards.

**Figure 1:**
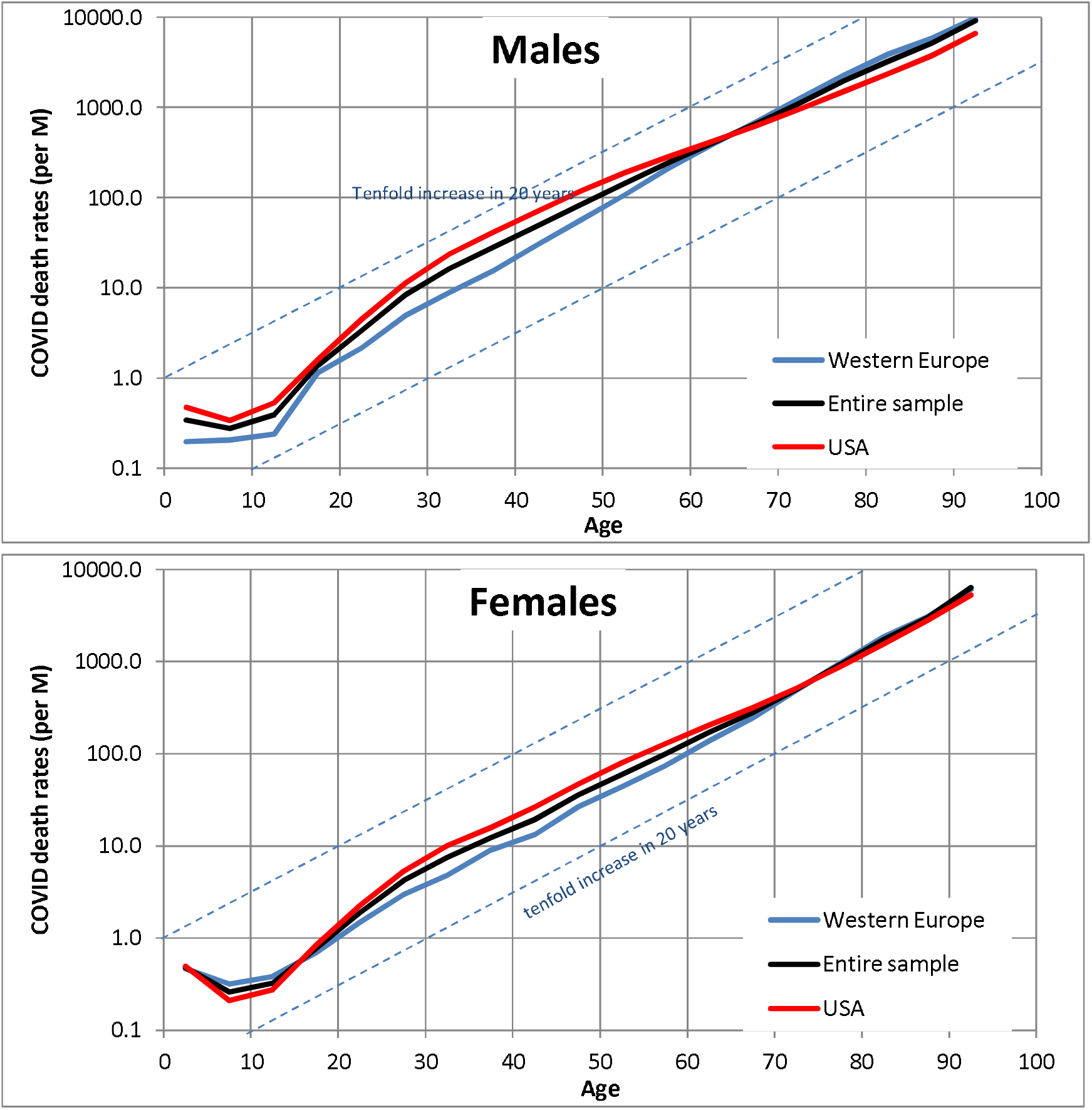
COVID-19 death rates by age and sex in death per million inhabitants, May 15, 2020. Notes: see Appendix 2 for detail on methods used

The different series follow an almost parallel pattern of sustained growth in death rates, from age 15 onward. The age gradient of COVID-19 mortality is pronounced, and death rates double for every six years of age. In fact, the charts show that death rates tend to increase tenfold for every 20 years of age (Table 1). Among all the women in the sample, COVID-19 mortality rates increase from a low 1.4 per million, at age 20, to 14 at 40, 127 at 60 and finally 1388 at 80. The other samples showed similar trends in female and male mortality.

**Table 1:**
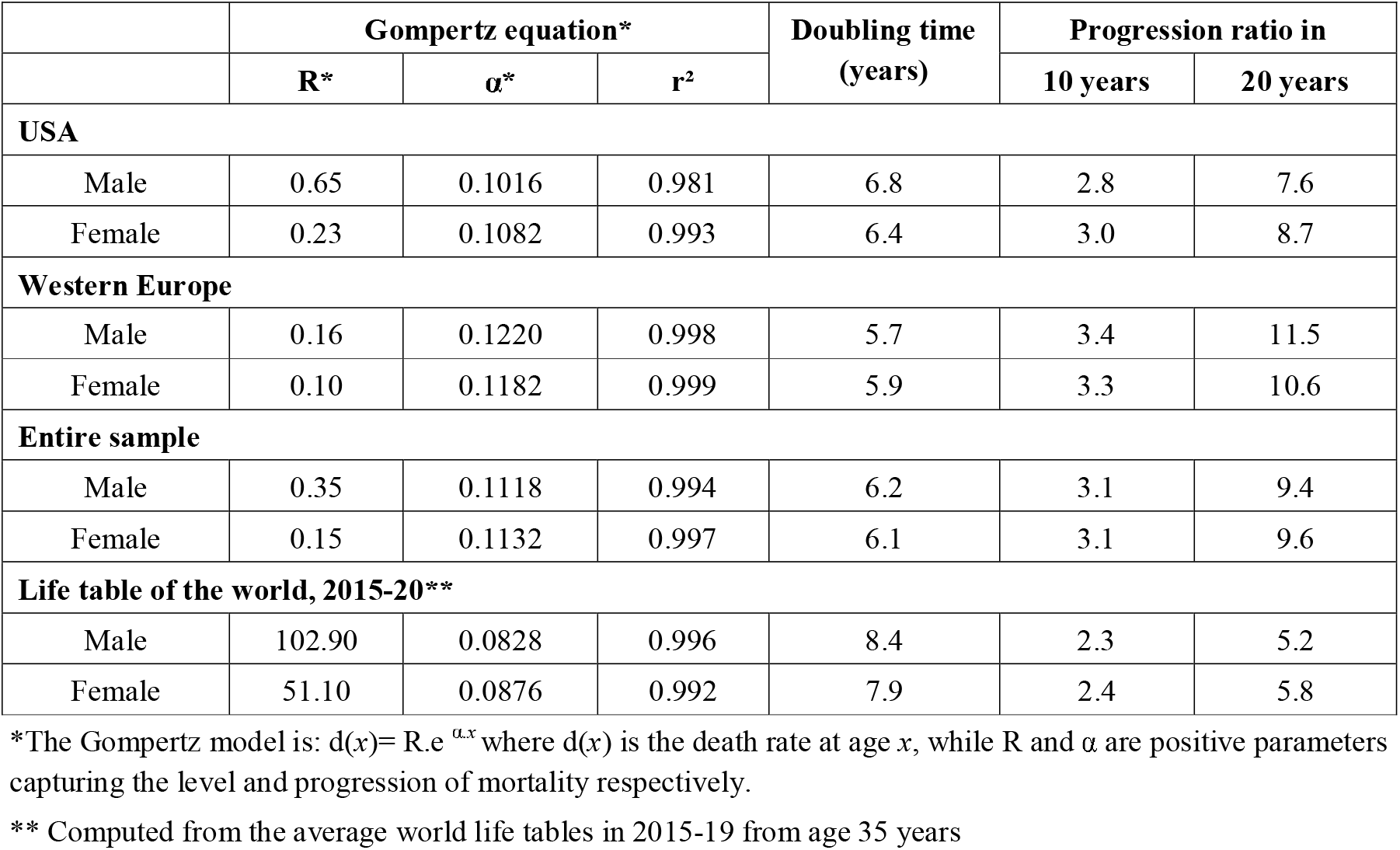
Age gradient of COVID-19 death rates and of the world’s average life tables for 2015-20.

|  | Gompertz equation* |  |  | Doubling time<br>(years) | Progression ratio in |  |
| --- | --- | --- | --- | --- | --- | --- |
| | R* | $\alpha^*$ | r<sup>2</sup> | | 10 years | 20 years |
| USA |  |  |  |  |  |  |
| Male | 0.65 | 0.1016 | 0.981 | 6.8 | 2.8 | 7.6 |
| Female | 0.23 | 0.1082 | 0.993 | 6.4 | 3.0 | 8.7 |
| Western Europe |  |  |  |  |  |  |
| Male | 0.16 | 0.1220 | 0.998 | 5.7 | 3.4 | 11.5 |
| Female | 0.10 | 0.1182 | 0.999 | 5.9 | 3.3 | 10.6 |
| Entire sample |  |  |  |  |  |  |
| Male | 0.35 | 0.1118 | 0.994 | 6.2 | 3.1 | 9.4 |
| Female | 0.15 | 0.1132 | 0.997 | 6.1 | 3.1 | 9.6 |
| Life table of the world, 2015-20** |  |  |  |  |  |  |
| Male | 102.90 | 0.0828 | 0.996 | 8.4 | 2.3 | 5.2 |
| Female | 51.10 | 0.0876 | 0.992 | 7.9 | 2.4 | 5.8 |

\*The Gompertz model is: $d(x) = R \cdot e^{\alpha \cdot x}$ where $d(x)$ is the death rate at age $x$ , while $R$ and $\alpha$ are positive parameters capturing the level and progression of mortality respectively.
\*\* Computed from the average world life tables in 2015-19 from age 35 years

In comparison, general mortality increased more gradually: from age 35, death rates from the world life table for 2015–20 increased by a factor of 5.5 for every 20 years of age. These steeper age gradients of COVID-19 mortality are typical of low-mortality settings, such as those found in industrialized countries, from which our death sample is drawn. Age-specific COVID-19 death rates may be less steep in moderate- or high-mortality countries, such as Asia, sub-Saharan Africa and Latin America, for which we currently have no disaggregated age data.

The extreme regularity of the increase in death rates by age, in all our samples after age 15, corresponds to an exponential growth of age-specific rates. COVID-19 death rates follow a typical mortality schedule, akin to the standard Gompertz equations used in adult mortality analysis (an alternative Weibull model proved less accurate).^15^ The results of the regressions, shown in Table 1, point to very high correlation levels (r^2^> .98) from age 15 to 90+. However, when we examined these more closely, we found that the age gradient of COVID mortality was slightly more pronounced in Western Europe, with death rates progressing faster there, with age, than in the USA.

### 3.2. Sex Differentials

Using these death rates, we can also compute the sex differentials of COVID-19 mortality. Figure 2 displays the sex ratio of mortality rates, computed as the ratio of male to female death rates by ageCZ Guilmoto: COVID-19 death rates by age and sex and the resulting mortality vulnerability group. We have also added the sex ratio of mortality for the life tables of the entire world for 2015–20.^13^

**Figure 2:**
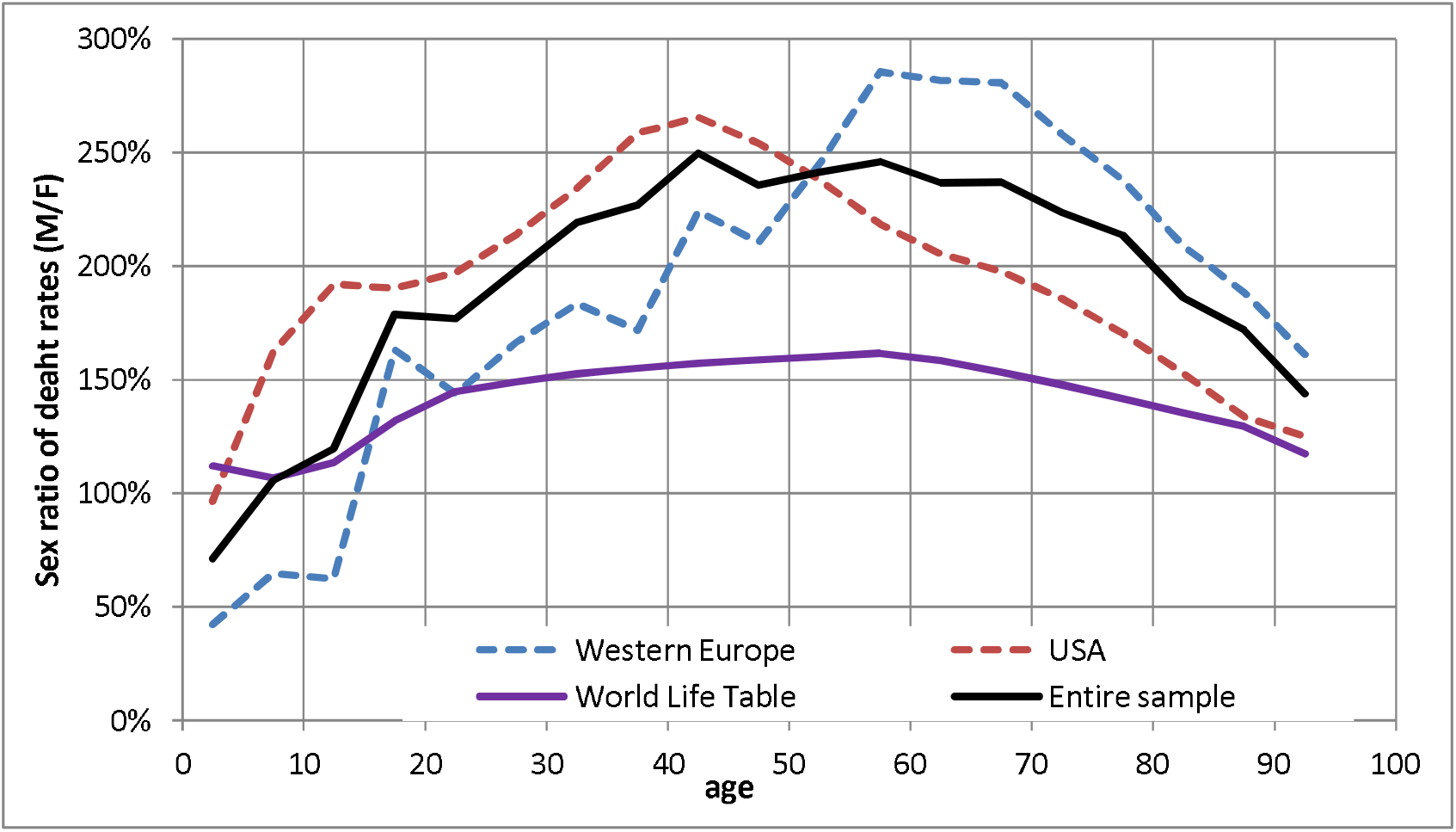
Sex ratio of COVID-19 death rates by sex of world mortality death rates in 2015-20.

It is well accepted that mortality is lower for females of all ages in most countries of the world. The world life table illustrates that the sex ratio of death rates is indeed greater than 1, and it gradually increases among adults, peaking at 1.6 before age 60. A comparison of the sex ratio of ordinary mortality and that of COVID-19 death rates, computed from our samples, demonstrates similar age schedules of mortality differentials. However, our comparison of series, in Figure 2, also emphasizes the presence of sex differentials that are unique to COVID-19 mortality.

The sex ratio of COVID-19 death rates increases rapidly; from a low ratio among the youth close to one, when death rates are infinitesimal, to a ratio of 2.5 at age 40-4 and 55–59. It gradually declines from this extreme level of excess male mortality, as men grow older, but it remains above 1.4 for the oldest age group of our series. Overall, the sex ratio of mortality appears considerably skewed— a feature that may not be explained merely by the different immune systems of adult men and women^16^.

The sex ratio of COVID-19 mortality differs between two samples. In the USA, the male mortality disadvantage tends to peak much earlier among 40–44-year-olds, compared to Western Europe, where it culminates among adults in their late fifties. The difference between mortality sex ratios, in the two samples, may be related to age at infection and to other unknown vulnerability factors linked to the case-fatality rates, which are specific to each region.^12,17–19^

### 3.3. Impact of age and Sex Structure on Mortality

This section examines the impact of demographic structures on potential COVID-19 mortality with the help of the COVID-19 structural vulnerability index (CSVI, see section 2). Table 2 presents the results of this comparative exercise for regions of the world. Regional variations in the COVID-19 structural vulnerability index are extremely pronounced. CSVI ranges from 28%, in Middle and Western Africa, to 260% in Western and Southern Europe. This indicates that, given similar age and sex patterns of COVID-19 mortality, countries in Western and Southern Europe would record almost ten times more deaths, proportionally, than in Western or Middle Africa. Variations of similar magnitude can also be computed by using the World Bank’s income levels, with the richest countries being almost seven times more vulnerable to COVID-19 mortality than less developed countries.

**Table 2:** COVID-19 structural vulnerability index by world regions (see text for definition)

| **Region** | **CSVI\*** |
| --- | --- |
| **World** | 100% |
| **High-income countries** | 218% |
| **Middle-income countries** | 83% |
| Upper-middle-income countries | 109% |
| Lower-middle-income countries | 61% |
| **Low-income countries** | 33% |
| **Sub-Saharan Africa** | 30% |
| Eastern Africa | 30% |
| Middle Africa | 28% |
| Southern Africa | 50% |
| Western Africa | 28% |
| **Northern Africa and Western Asia** | 62% |
| Northern Africa | 59% |
| Western Asia | 64% |
| **Central and Southern Asia** | 64% |
| Central Asia | 57% |
| Southern Asia | 64% |
| **Eastern and South-eastern Asia** | 119% |
| Eastern Asia | 136% |
| South-eastern Asia | 76% |
| **Australia/New Zealand** | 191% |
| **Oceania (excluding Australia and New Zealand)** | 42% |
| **Europe** | 219% |
| Eastern Europe | 170% |
| Northern Europe | 223% |
| Southern Europe | 264% |
| Western Europe | 257% |
| **Northern America** | 192% |
| **Latin America and the Caribbean** | 100% |
| Caribbean | 121% |
| Central America | 83% |
| South America | 104% |

\*: The CSVI index is computed as the ratio of potential COVID-19 death rates in various regions to the CSVI for the entire world.
Rates for individual countries are available from the author

A separate analysis of the impact of age structures by sex shows that age composition matters much more to COVID mortality than sex composition (not reported here). This is due to the fact that age structures are much more heterogeneous across the world, because of the large variations in fertility levels, which have been observed in the past thirty years, and their direct impact on the proportion of elderly persons in populations.

The CSVI of individual countries is shown on the world map, in Figure 3. Several large countries, such as Brazil, China, Mexico and Turkey, are in an intermediate situation, with a vulnerability index to COVID-19 similar to the world average. However, the index ranges more widely across the world than across regions, ranging from 25% to 355%. The countries with lowest indexes include Uganda (20%), Zambia, Angola, Burkina Faso, and Mali (all <25%). The countries with the lowest vulnerability index, outside sub-Saharan Africa, include Afghanistan (27%), Yemen and the United Arab Emirates (30%).

**Figure 3:**
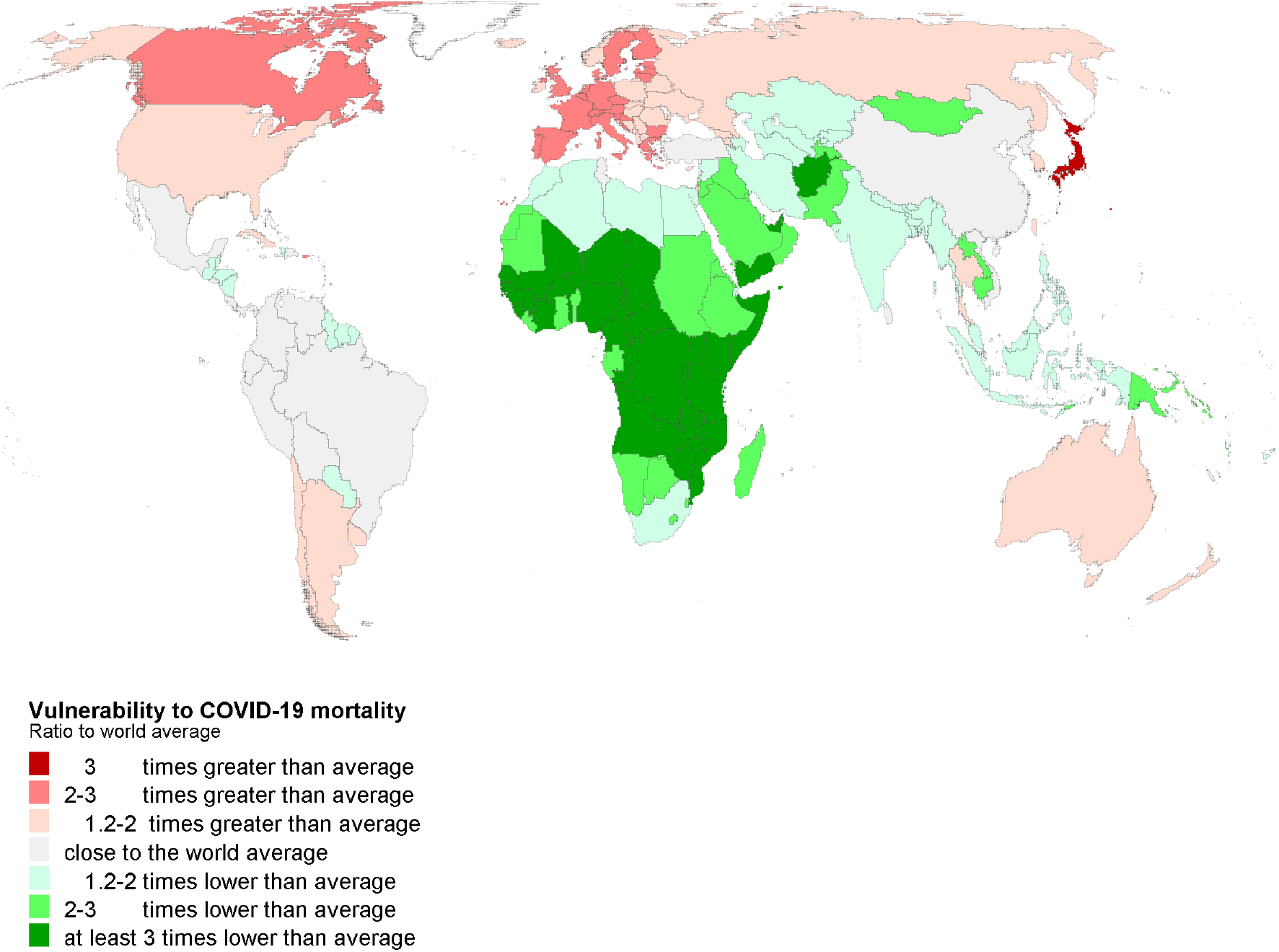
OVID-19 structural vulnerability index (CSVI) of individual countries in 2020. (see text for definition)

In contrast, the countries that are potentially the most exposed to COVID mortality have indices above 250% and include France, Spain, Portugal, Germany, Greece and Italy in Western and Southern Europe. The highest value is observed in Japan (355%). The ratio between the least and most vulnerable countries is in a proportion of one to 17. This suggests that an overall death rate from COVID-19 would be 17 times lower in Uganda than in Japan, if both countries were to experience the same age and sex-specific death rates, when computed on our sample.

The vulnerability index tends to be especially high in those countries that have been most affected by COVID-19 mortality, such as Italy or Belgium.(8,13) The CSVI emerges in fact as a powerful predictor of COVID-19 mortality rates. As Figure 4 indicates, higher levels of COVID-19 vulnerability are associated with higher COVID-19 death rate among 188 countries for which we had both indicators. The correlation coefficient (*r*^2^) between CSVI and current COVID-19 death rates (log transformed) at country level is as high as .44. This suggests that almost half of the variations observed today in COVID-19 mortality across the world can be attributed to the specific age and sex structures of affected countries.

**Figure 4:**
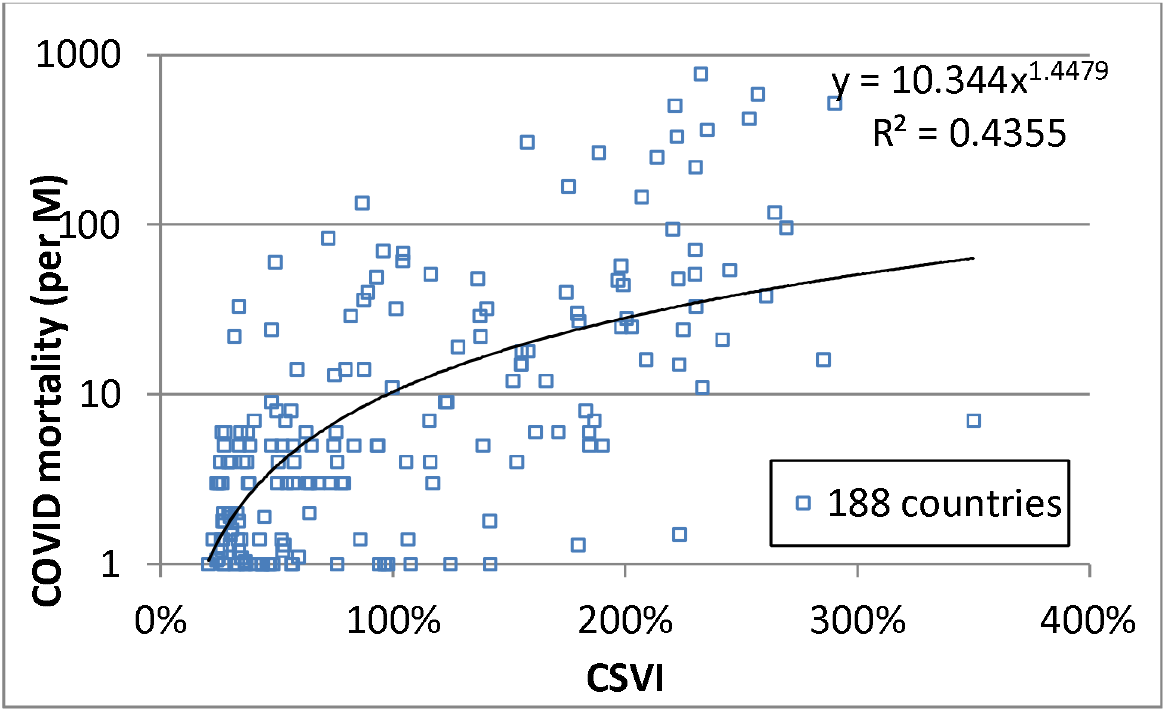
COVID-19 structural vulnerability index (CSVI) and current mortality rates of individual countries. (see text for definition)

We may, however, notice that the correspondence between potential vulnerability and actual COVID-19 mortality is far from perfect. Some countries, which are highly vulnerable to the pandemic, such as Japan, Greece or Bulgaria, have remained almost untouched by COVID-19 mortality so far. In contrast, some hard-hit countries, such as Iran, Ecuador or Peru, display CSVI levels that are below 1. Many other factors have contributed to COVID-19 mortality, observed so far, such as the date of the onset of the pandemic, public health infrastructures and the policies introduced to combat the virus, population density and mobility, and the magnitude of exchanges with the main core infection zones.

## 4. Discussion and Conclusion

The recent publication of disaggregated information on COVID-related mortality from western countries represents a breakthrough in the demographic analysis of COVID mortality.^7^ It permits the comparative study of its potential impact, by both age and gender, on populations of different structures. In this paper we have assembled the largest database available today, which is based on the age and sex characteristics of 183,619 deaths, attributed to COVID-19. These deaths occurred since the beginning of 2020, in nine different countries, and represent 61% of all deaths attributed to COVID-19 today.

Our analysis of this unique dataset shows the regularity of the distribution of COVID deaths and their age and sex patterns of COVID mortality in industrialized countries. Incomplete coverage and the unfinished character of the pandemic have directly and undeniably affected the intensity of mortality and, consequently, the level of absolute death rates has not been highlighted herein. In spite of these potential shortcomings, we can already draw several lessons from our analysis:

- Reported cases of deaths, below age 15, are too few to allow an estimation of COVID-19 mortality below this age.
- COVID death rates increase with extremely regularly from age 15 to 90+ and follow a typical exponential Gompertz model, with rates doubling every six years.
- The age gradient of COVID mortality is steeper than that of ordinary mortality, causing a greater concentration of deaths in old age.
- Sex differentials in COVID mortality are unusually pronounced, with the mortality rates of men aged 25–80 being two times greater than those of women.
- The considerable variations in the potential impact of COVID mortality, between world regions, in relation to their demographic structures.
- Individual countries vary in their vulnerability to COVID mortality by a factor of one to 17 because of the proportion of elderly persons in their populations.
- The COVID-19 vulnerability index accounts for 46% of the variations in current COVID-19 mortality observed across the world.

Our analysis has also highlighted new questions that require in-depth statistical, clinical and epidemiological attention. We suggest a non-exhaustive list of such unresolved issues, as follows.

- The factors behind the steeper age gradient of mortality in Western Europe
- The potential bias in demographic estimation, because of the better registration of COVID deaths that follow hospitalization as compared to deaths in old age homes or at home.
- The nature of factors accounting for the pronounced excess COVID mortality observed among men at all ages.
- The similarity of age schedules of COVID mortality in countries and regions not sampled here, such as China, India, Iran, Latin America, and Eastern Europe.

A final caveat is that our analysis relies on datasets that suffer from two main potential limitations: the under-registration of the various forms of COVID-related excess mortality and the non-availability of disaggregated demographic information for many countries outside Western Europe and the USA. We hope that national statistical offices, from other countries affected by COVID mortality, will soon enrich the demographic sources used in this paper.

## Data Availability

original data available from Internet sources

## Appendix 1: Data sources

**Table 3:**
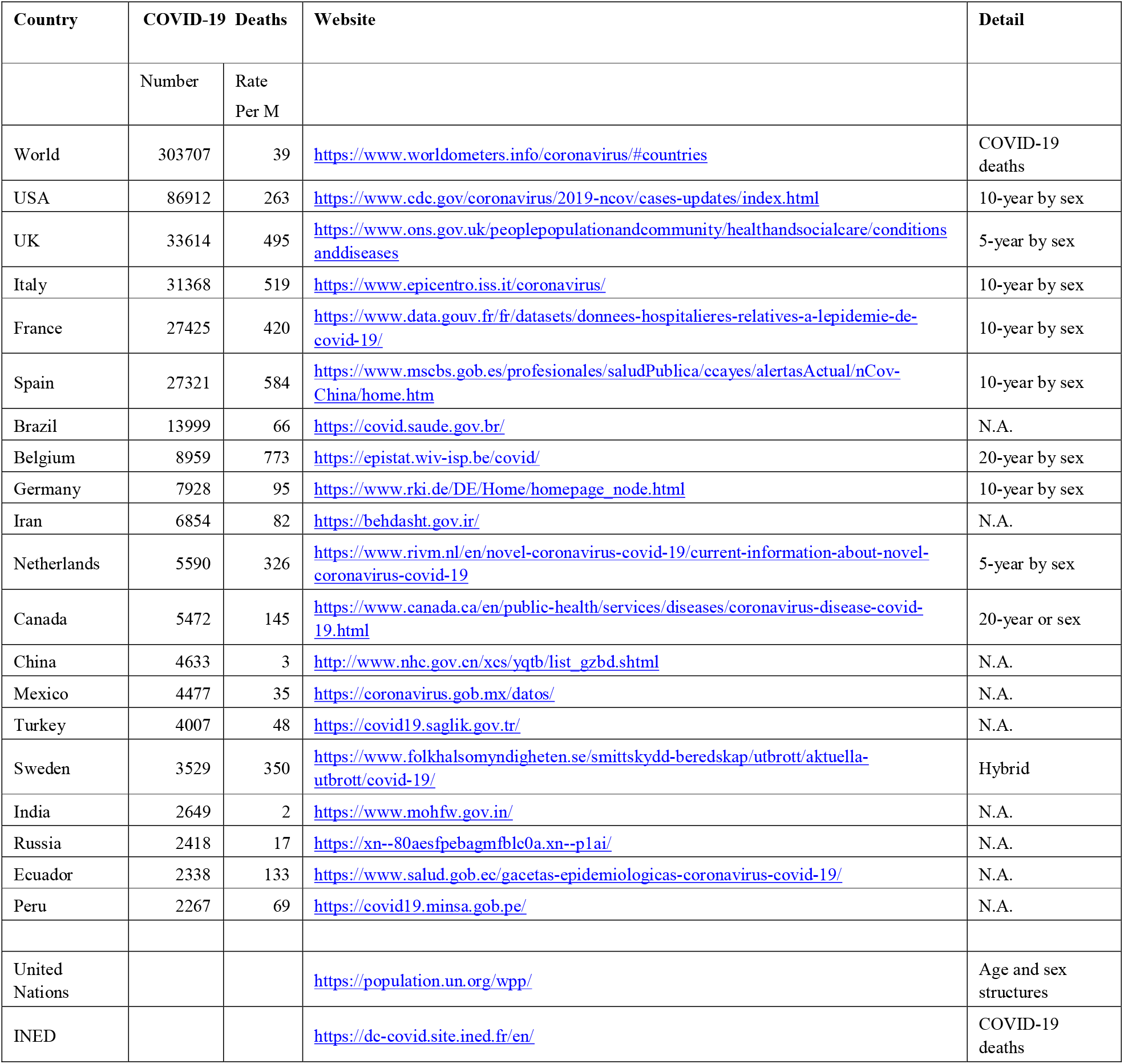
Data sources for population structures and for deaths in countries with more than 2000 COVID-19 deaths.

CZ Guilmoto: COVID-19 death rates by age and sex and the resulting mortality vulnerability

## Appendix 2 Standardization of age and sex-specific COVID-19 rates

The age and sex classifications of original COVID-19 deaths vary greatly by country. Hereunder, we describe the original data format of each country and the methods this study used to reclassify the deaths, by age and sex, into five-year deaths by sex.

England, Wales and the Netherlands:

- deaths already available by 5-year age groups and sex

USA, Italy, Spain, France, and Germany:

- deaths are available by 10-year age groups and sex

- rates by 5-year age groups and sex were estimated by computing the geometric mean of rates of adjacent ages
- estimated deaths by 5-year age groups and sex were computed from rates, and then standardized to tally observed death totals by sex.

Belgium

- deaths are available by 20-year age group and sex

- deaths were redistributed into 10-year groups, using ratios deduced from computed European COVID-19 rates
- the same procedure as described for the USA, Italy, Spain, France, and Germany was then followed to estimate the 5-year rates.

Sweden

- Deaths are available in a hybrid age format (5-, 10-year and larger age groups).

- Deaths by sex, in broader age groups, were redistributed into five-year age groups using the proportions computed for the Netherlands

The comparative number of deaths and deaths rates are taken from estimates given by Worldometer on May 15, 2020, to compute the coverage rate of age and sex statistics.^10^.

Deaths for each country were reweighted to adjust for variations in coverage (reweighted deaths= sample deaths/coverage rate).

## Appendix 3: COVID-19 death rates in the nine sample countries

**Figure 5:**
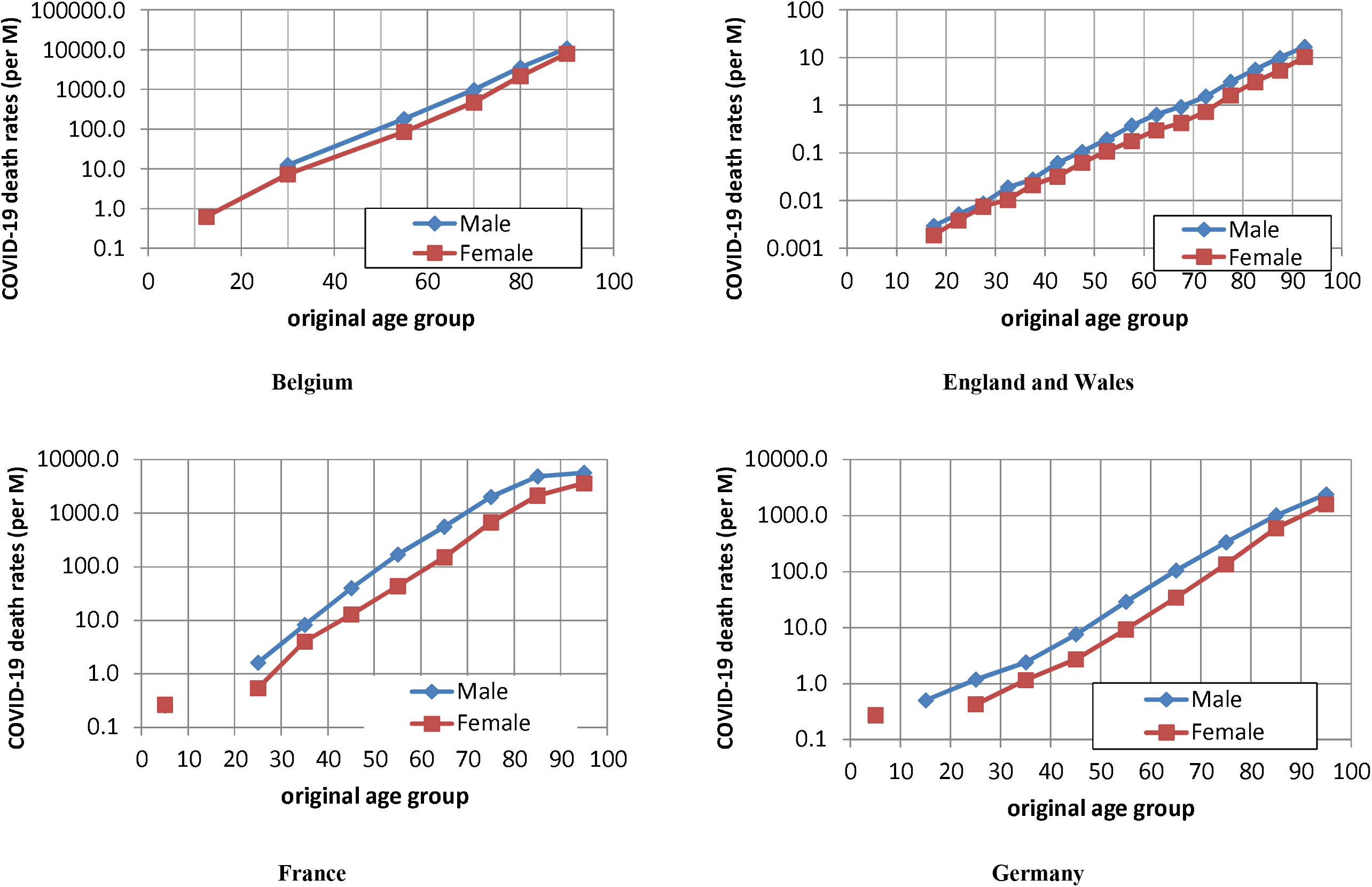
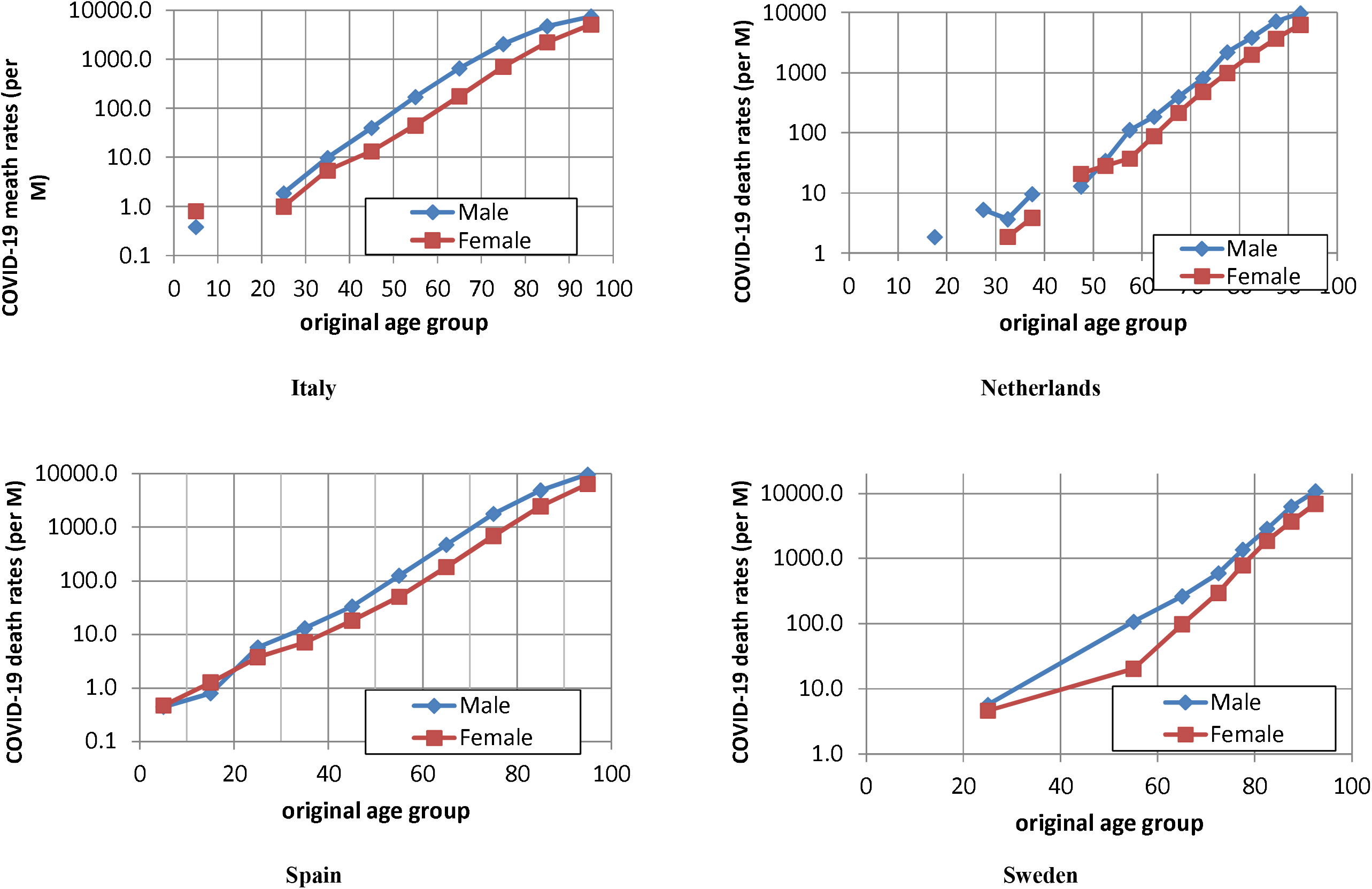
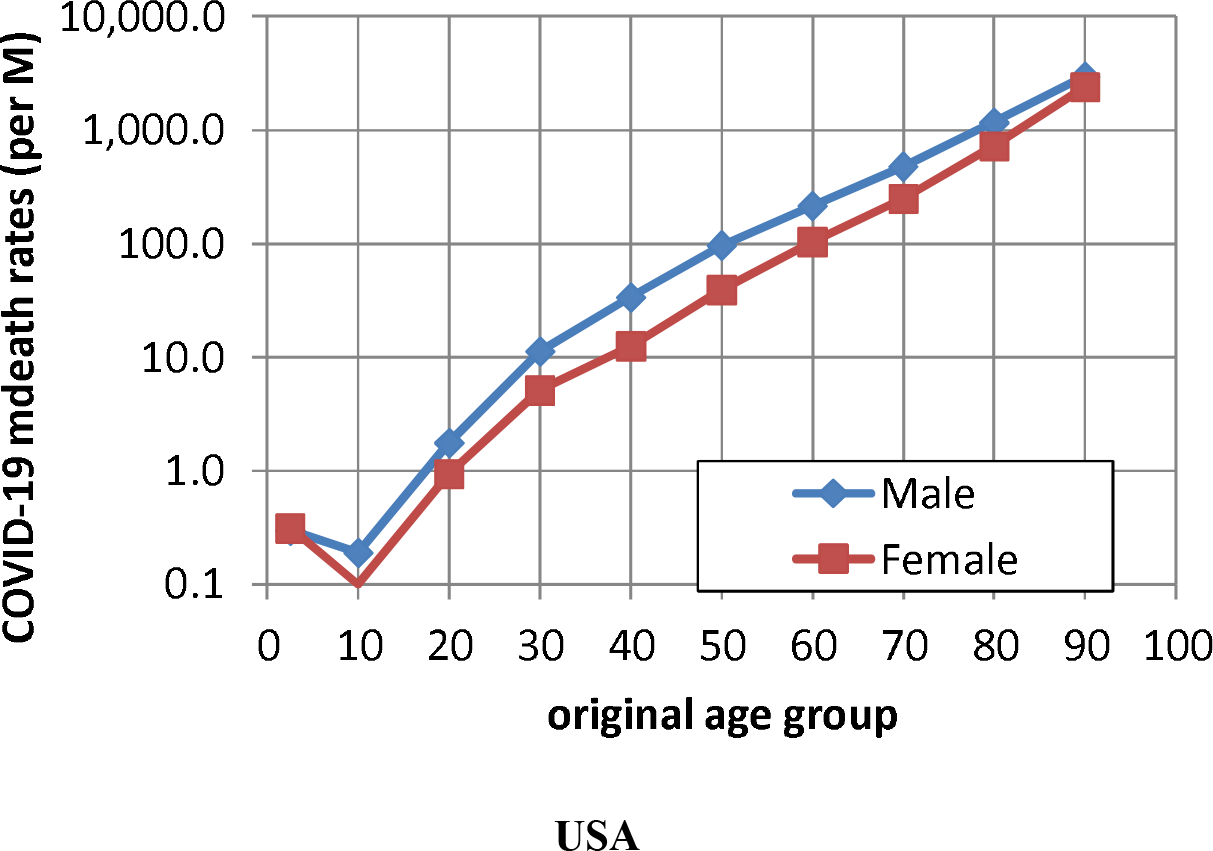
COVID-19 death rates by age and sex per million people based on original death data, May 15, 2020.

- Age format varies (see Appendix 2)
- Computed on raw death counts
- Rates below. 1 per million not shown
- Rates shown at the middle of the age group (e.g. 12.5 for the 0–24 years)

## Appendix 4: estimation of age- and sex-specific COVID-19 death rates in three samples.

**Table 4:** COVID-19 death rates per million people by age group and sex, USA, Western Europe and entire sample, 15 May 2020.

|  | USA |  |  | Western Europe |  |  | Entire sample |  |  |
| --- | --- | --- | --- | --- | --- | --- | --- | --- | --- |
|  | Male | Female | M/F* | Male | Female | M/F* | Male | Female | M/F* |
| **0–4** | 0 | 0 | 96% | 0 | 0 | 42% | 0 | 0 | 71% |
| **5–9** | 0 | 0 | 162% | 0 | 0 | 65% | 0 | 0 | 106% |
| **10–14** | 1 | 0 | 192% | 0 | 0 | 62% | 0 | 0 | 120% |
| **15–19** | 2 | 1 | 190% | 1 | 1 | 163% | 1 | 1 | 179% |
| **20–24** | 4 | 2 | 197% | 2 | 1 | 144% | 3 | 2 | 177% |
| **25–29** | 11 | 5 | 214% | 5 | 3 | 167% | 8 | 4 | 198% |
| **30–34** | 24 | 10 | 234% | 9 | 5 | 184% | 16 | 7 | 219% |
| **35–39** | 41 | 16 | 259% | 15 | 9 | 172% | 28 | 12 | 227% |
| **40–44** | 70 | 26 | 266% | 30 | 13 | 224% | 48 | 19 | 250% |
| **45–49** | 119 | 47 | 254% | 56 | 27 | 211% | 84 | 36 | 236% |
| **50–54** | 189 | 79 | 238% | 107 | 44 | 245% | 143 | 59 | 242% |
| **55–59** | 282 | 129 | 219% | 211 | 74 | 286% | 244 | 99 | 246% |
| **60–64** | 420 | 205 | 205% | 389 | 138 | 282% | 404 | 171 | 237% |
| **65–69** | 626 | 317 | 198% | 691 | 246 | 281% | 661 | 279 | 237% |
| **70–74** | 956 | 515 | 186% | 1264 | 489 | 258% | 1121 | 501 | 224% |
| **75–79** | 1494 | 876 | 170% | 2281 | 958 | 238% | 1977 | 925 | 214% |
| **80–84** | 2357 | 1543 | 153% | 3869 | 1852 | 209% | 3224 | 1730 | 186% |
| **85–89** | 3756 | 2811 | 134% | 5798 | 3076 | 188% | 5136 | 2991 | 172% |
| **90+** | 6570 | 5254 | 125% | 9786 | 6077 | 161% | 9168 | 6382 | 144% |
| *M/F: sex ratio of age specific mortality rates (in %)<br>Rates computed on deaths adjusted for national coverage level (see Appendix 2)<br>All rates given per million people. |  |  |  |  |  |  |  |  |  |

## Notes

### Competing Interest Statement

The authors have declared no competing interest.

### Funding Statement

no funding

